# CSF p-tau Predicts Asymmetric Hippocampal Atrophy in Cognitively Unimpaired Older Adults

**DOI:** 10.64898/2026.05.04.26352391

**Authors:** Elham Ghanbarian, Lukai Zheng, Tianchen Qian, Crystal M. Glover, S. Ahmad Sajjadi, Maria M. Corrada, Joshua D. Grill, Alzheimer’s Disease Neuroimaging Initiative, Ali Ezzati

**Affiliations:** Department of Neurology, University of California, Irvine, CA, USA; Center for Design, Discovery, and Data Analytics (3D Center) in Neurological Disorders, University of California, Irvine, CA, USA; Department of Statistics, University of California, Irvine, CA, USA; Alzheimer’s Disease Research Center, Institute for Memory Impairments and Neurological Disorders (UCI MIND), University of California, Irvine, CA, USA; Department of Psychiatry and Human Behavior, University of California, Irvine, CA, USA; Department of Epidemiology and Biostatistics, University of California, Irvine, CA, USA

**Keywords:** Alzheimer’s disease, Hippocampal asymmetry, Hippocampal atrophy, Phosphorylated tau 181 (p-tau181), Amyloid-β 42 (Aβ42)

## Abstract

**Background:** Hippocampal atrophy is a core marker of neurodegeneration in dementia, particularly in Alzheimer’s disease (AD). However, most studies focus on total hippocampal volume loss and overlook hemispheric asymmetry, which may reflect distinct biological processes. While tau pathology is closely linked to medial temporal lobe degeneration, it remains unclear whether tau is associated with asymmetric patterns of hippocampal atrophy.

**Methods:** We analyzed 483 cognitively unimpaired participants from the Alzheimer’s Disease Neuroimaging Initiative (ADNI) with baseline cerebrospinal fluid (CSF) phosphorylated tau (p-tau181) and amyloid-β (Aβ42) measurements and longitudinal structural MRI data over 10 years of follow-up. Total hippocampal volume and hemispheric asymmetry, defined as the absolute difference between left and right hippocampal volumes (|L−R|), were quantified at each visit. Linear mixed-effects models assessed associations between baseline CSF biomarkers and longitudinal changes in hippocampal asymmetry, adjusting for demographic factors, APOE ε4 status, and baseline hippocampal volume.

**Results:** Higher baseline CSF p-tau181 was associated with greater increases in hippocampal asymmetry over time (β = 1.20, SE = 0.43, p = 0.006). This association remained significant after additional adjustment for total hippocampal volume and baseline CSF Aβ42. CSF total tau was highly correlated with p-tau181 (Spearman’s ρ =0.98, p < 0.001) and showed comparable associations with hippocampal measures. In contrast, baseline Aβ42 was not associated with subsequent changes in hippocampal asymmetry. Both p-tau181 and Aβ42 were associated with faster decline in total hippocampal volume. In amyloid-stratified analyses, p-tau181 was associated with increasing hippocampal asymmetry only among amyloid-negative individuals, whereas its association with total hippocampal atrophy was observed primarily in amyloid-positive individuals.

**Conclusions:** CSF p-tau181 is associated not only with overall hippocampal atrophy but also with progressive hemispheric asymmetry, suggesting that tau-related neurodegeneration may manifest as both magnitude and imbalance of tissue loss. These findings support hippocampal asymmetry as a complementary neuroimaging marker that may capture non-amyloid-related medial temporal lobe degeneration in cognitively unimpaired older adults.

## 1. Introduction

Hippocampal atrophy is one of the most robust and widely used neuroimaging markers of Alzheimer’s disease (AD), reflecting neurodegeneration across both preclinical and clinical stages.^1^ Progressive volume loss within medial temporal lobe (MTL) structures, particularly the hippocampus and entorhinal cortex, closely tracks cognitive decline and risk of progression to dementia.^2^ In addition, MRI-derived measures of hippocampal atrophy have been consistently shown to correlate with underlying neuropathology, supporting its role as valid in-vivo biomarkers of disease progression.^3^

Accumulating evidence suggests that tau pathology, rather than amyloid burden, is more directly linked to neurodegeneration and atrophy in AD.^4, 5^ Tau deposition begins within MTL structures, and regional atrophy closely mirrors the pattern of tau spread in these regions, whereas amyloid accumulation does not show the same tight coupling with atrophy.^4, 6^ In preclinical AD, accelerated longitudinal atrophy of MTL, including hippocampus, has been reported in individuals with elevated tau, supporting that hippocampal atrophy emerges in the context of tau pathology.^5, 7^ In particular, post-mortem studies have demonstrated that phosphorylated tau deposition is strongly associated with hippocampal atrophy.^8, 9^

While overall hippocampal atrophy is well established as a diagnostic and prognostic marker of AD, most prior work has treated total hippocampal volume as the primary structural summary of disease-related degeneration. However, total volume as a bilateral aggregate measure may obscure an important feature of MTL neurodegeneration, which is its lateralization. If degeneration does not occur symmetrically across the two hippocampi, then the overall magnitude of volume loss may fail to capture asymmetric atrophy, which may represent a biologically meaningful pattern.

Prior work on hippocampal asymmetry in AD has been mixed, suggesting that asymmetry is not simply a universal feature of AD severity but may instead reflect specific biological subtypes or pathological heterogeneity.^10–14^ Recent work from our group has shown that while total hippocampal volume is associated with cognitive performance in multiple domains, greater left-right hippocampal asymmetry is primarily associated with accelerated memory decline, especially among amyloid-negative individuals.^15^ These findings suggest that hippocampal left-right asymmetry may provide complementary information beyond total hippocampal volume and capture non-amyloid neurodegenerative pathways. Furthermore, conditions not primarily defined by amyloid pathology, such as hippocampal sclerosis of aging (HSA), often associated with limbic-predominant age-related TDP-43 encephalopathy (LATE) neuropathological changes, can produce marked asymmetric hippocampal atrophy.^16, 17^

Although the role of tau pathology in MTL and particularly hippocampal atrophy has been well demonstrated, it remains unclear whether tau is associated primarily with global hippocampal volume loss, or whether it also contributes to asymmetric patterns of neurodegeneration over time. This distinction is important because total atrophy and asymmetric atrophy may index distinct processes as one reflects overall tissue loss and the other reflects selective or lateralized vulnerability within the hippocampus. If tau is associated with progressive asymmetry independent of total volume, this would suggest that tau-related neurodegeneration reflects not only the magnitude of tissue loss but also its hemispheric distribution,^18^ with important clinical implications.

In this study, we investigated whether baseline cerebrospinal fluid (CSF) phosphorylated tau 181 (p-tau181) is associated with longitudinal changes in total hippocampal atrophy and hemispheric asymmetry in cognitively unimpaired individuals. By focusing on this population, we aimed to determine whether p-tau181 is associated with early, subtle alterations in hippocampal structure prior to the onset of significant atrophy and downstream cognitive impairment. Specifically, we examined whether elevated CSF p-tau181 is associated with increasing hippocampal asymmetry over time, independent of total hippocampal volume, and whether these relationships differ by amyloid burden. By separating the magnitude of hippocampal atrophy from hemispheric imbalance, this study aims to determine whether tau-related neurodegeneration in cognitively unimpaired older adults is expressed primarily as global tissue loss, asymmetric degeneration, or both.

## 2. Methods

### 2.1. Participants

#### 2.1.1. ADNI study design

Data used in the preparation of this article were obtained from the Alzheimer’s Disease Neuroimaging Initiative (ADNI) database (adni.loni.usc.edu). The ADNI was launched in 2003 as a public-private partnership, led by Principal Investigator Michael W. Weiner, MD. The primary goal of ADNI has been to test whether serial magnetic resonance imaging (MRI), positron emission tomography (PET), other biological markers, and clinical and neuropsychological assessment can be combined to measure the progression of mild cognitive impairment (MCI) and early AD. Importantly, ADNI diagnoses are based on standardized clinical criteria and do not require pathological confirmation, allowing for etiologic heterogeneity at the clinical endpoint. ADNI data collection was approved by the institutional review boards of all participating institutions and informed written consent was obtained from all participants. Detailed information on measures and methods of assessment in the ADNI project are available at http://www.adni.loni.usc.edu.

#### 2.1.2. Study participants

We included cognitively unimpaired participants from ADNI (ADNI 1, ADNI GO, ADNI 2, and ADNI 3) who met the following criteria at baseline: (1) availability of at least two longitudinal T1-weighted MRI processed through the ADNI FreeSurfer pipeline and (2) availability of baseline CSF concentrations of amyloid-β (Aβ42) and phosphorylated tau at threonine 181 (p-tau-181). Participants with a baseline diagnosis of mild cognitive impairment and dementia were excluded. After applying these criteria 483 cognitively unimpaired (CU) participants were included in the study.

In the ADNI study, diagnostic classification at baseline followed standardized criteria. CU individuals were required to have a Mini-Mental-State Examination (MMSE) scores of 24 or higher; Clinical Dementia Rating (CDR) score of 0, and no evidence of depression.

### 2.2. Study measures and outcomes

#### 2.2.1. MRI and hippocampal asymmetry

Structural MRI data were preprocessed using FreeSurfer (http://surfer.nmr.mgh.harvard.edu/) by the ADNI MRI Core as part of the ADNI standard pipelinesset. FreeSurfer methods for identifying and calculation of regional brain volumes have been described in detail previously.^19^

For this study, left (L) and right (R) hippocampal volumes were adjusted for intracranial volume (ICV). Total hippocampal volume was defined as the sum of left and right hippocampal volumes (L+R), and hippocampal asymmetry as the absolute hemispheric difference (|L − R|).

Both total hippocampal volume and hemispheric asymmetry were included simultaneously in all statistical models to distinguish effects attributable to total atrophy from those reflecting asymmetric neurodegeneration. To assess potential collinearity between hippocampal volume and asymmetry, we calculated variance inflation factors (VIFs) and condition indices using a linear regression model including both variables. VIF values were < 5 and condition indices were < 30, indicating no evidence of problematic multicollinearity. Although absolute hemispheric difference may still scale with overall hippocampal volume, the absence of collinearity and the simultaneous modeling of total volume and asymmetry would allow for estimation of independent associations with cognitive and clinical outcomes.

#### 2.2.2. CSF Biomarkers

CSF samples were batch processed by the ADNI Biomarker Core at the University of Pennsylvania School of Medicine using Roche Elecsys assay.^20^ Baseline CSF p-tau181 and Aβ42□ levels were included as continuous variables in all models. For descriptive purposes and stratified models, amyloid positivity was defined as CSF Aβ42 < 977 pg/mL, and p-tau181 positivity was defined as CSF p-tau181 > 27 pg/mL, consistent with validated ADNI cutpoints.^21^. Additionally, baseline total tau (t-tau) was analyzed as a continuous variable for comparison with p-tau181.

#### 2.2.3. Tau PET Imaging

Tau PET ([18F]flortaucipir, AV-1451) data were obtained from the ADNI database in fully preprocessed form using the standardized ADNI pipeline. T1-weighted MRI scans were processed with FreeSurfer (v5.3) to define regions of interest (ROIs), and PET images were co-registered to the corresponding MRI using SPM5. SUVR values were calculated by normalizing regional tracer uptake to inferior cerebellar gray matter. A temporal meta-ROI was defined as the volume-weighted average of bilateral entorhinal, amygdala, fusiform, inferior temporal, and middle temporal regions. Tau positivity was determined using a previously validated SUVR threshold of ≥1.37 for the meta-ROI.^22^

### 2.3. Statistical analysis

Differences in participant characteristics across sex and age subgroups were tested using independent-sample t tests for continuous variables and chi-squared test for categorical variables. Associations between CSF p-tau181 and t-tau, as well as between these biomarkers and tau PET SUVR were assessed using Spearman rank correlation. Additionally, partial correlation adjusted for age, sex and amyloid burden were used to evaluate the associations between CSF tau biomarkers and tau PET.

To evaluate the associations between baseline p-tau181 and Aβ42 and longitudinal changes in total hippocampal volume and asymmetry, we used linear mixed-effects (LME) models. Time was modeled as a continuous variable representing years since baseline visit. The base model included fixed effects for demographics (age, sex, years of education), APOE ε4 status, time, baseline p-tau181and its interactions with time. When evaluating the hippocampal asymmetry as an outcome, the models were also adjusted for the baseline total hippocampal volume. Random intercepts and slopes for time were included at the participant level to capture individual variability in baseline hippocampal asymmetry and its rate of change. Separate models were fit for hippocampal asymmetry and total volume. The estimates for p-tau181 reflect the associations with baseline hippocampal asymmetry or baseline total volume (time 0), whereas the p-tau × time interactions reflect the associations with the rate of change in asymmetry over time or total volume over time.

In secondary models, baseline Aβ42 was added as a continuous variable to the base models to evaluate whether associations between p-tau181 and hippocampal measures were different when accounting for amyloid levels. In addition, amyloid-stratified analyses were conducted to examine potential effect modification by baseline binary amyloid status. Separate LME models were fit within amyloid-positive and amyloid-negative groups using identical model specifications.

To evaluate the associations between p-tau181 and longitudinal trajectories of left and right hippocampal volumes, we fitted separate LME models with left and right hippocampal volume as the respective outcomes. All statistical analyses were conducted in SPSS version 27 (IBM Corp, Armonk, NY, USA).

## 3. Results

### 3.1. Cohort characteristics

The study cohort consisted of 483 cognitively unimpaired individuals, of whom 275 (56.9%) were females, and the mean age was 72.9 ± 6.15 years. The average total hippocampal volume in the entire sample was 7.62 ± 0.87 cm^3^, and the average hippocampal asymmetry was 0.22 ± 0.17 cm^3^. On average, the right hippocampus was larger than the left in the entire cohort (3.87 ± 0.47 vs 3.75 ± 0.44 cm^3^, paired t-test, *p* < 0.001). Based on CSF p-tau181 levels, 378 individuals were classified as p-tau181 negative (p-tau181-) and 105 as p-tau181 positive (p-tau181+). At baseline, total hippocampal volume was smaller in p-tau181+ group (*p* = 0.041), but there was no significant difference in hippocampal asymmetry between p-tau181 negative and positive groups (*p* = 0.245). There was no significant difference in total hippocampal volume or hippocampal asymmetry between the amyloid-positive (Aβ+) and amyloid-negative (Aβ-) groups.

Demographic, biomarker, and cognitive domain characteristics of the study cohort are summarized in Table 1. Mean change from baseline in total hippocampal volume and hippocampal asymmetry across follow-up visits (10 years from baseline) in the overall cohort is presented in Fig 1.

**Table 1.** Participants’ demographics at baseline.

| Characteristic | All | p-tau181- | p-tau181+ | Aβ42- | Aβ42+ |
| --- | --- | --- | --- | --- | --- |
| Participants, N (%) | 483 | 378 (78.3) | 105 (21.7) | 314 (65.01) | 169 (34.99) |
| Female Sex, n (%) | 275 (56.9) | 214 (56.6) | 61 (58.1) | 179 (57) | 96 (56.8) |
| APOE ε4 carrier, n (%) | 151 (31.3) | 107 (28.3) | 44 (41.9) | 61 (19.4) | 90 (53.3) |
| Age, yr | 72.90 (6.15) | 72.19 (5.92) | 75.44 (6.30) | 72.52 (6.12) | 73.60 (6.15) |
| Education, yr | 16.53 (2.52) | 16.47 (2.53) | 16.75 (2.47) | 16.55 (2.54) | 16.50 (2.48) |
| CSF Aβ42, pg/mL | 1344.32 (664.97) | 1336.06 (586.10) | 1374.07 (896.09) | 1690.82 (565.32) | 700.54 (178.61) |
| CSF p-tau181, pg/mL | 21.46 (8.96) | 17.64 (4.49) | 35.23 (7.36) | 20.55 (7.63) | 23.16 (10.82) |
| CSF p-tau181/Aβ42 | 0.02 (0.02) | 0.02 (0.01) | 0.04 (0.02) | 0.01 (0.01) | 0.04 (0.02) |
| CSF total tau, pg/mL | 235.75 (86.1) | 199.56 (48.13) | 366.02 (63.80) | 233.59 (80.36) | 239.75 (95.98) |
| Tau PET SUVR, temporal meta-ROI* | 1.21 (0.15) | 1.18 (0.12) | 1.29 (0.21) | 1.18 (0.07) | 1.26 (0.22) |
| Total Hippocampal Volume, cm<sup>3</sup> | 7.62 (0.87) | 7.66 (0.88) | 7.46 (0.84) | 7.64 (0.85) | 7.59 (0.91) |
| Hippocampal asymmetry, cm<sup>3</sup> | 0.22 (0.17) | 0.22 (0.16) | 0.24 (0.18) | 0.22 (0.17) | 0.22 (0.17) |

Continuous variables are expressed as mean ± standard deviation (SD), and categorical variables as counts (percentages). Aβ 42, amyloid-beta 42; p-tau18, phosphorylated tau at threonine 181. Aβ 42 and p-tau181 status is defined by CSF biomarker cutoff. \* Tau PET data was available for 130 individuals.

**Figure 1.**
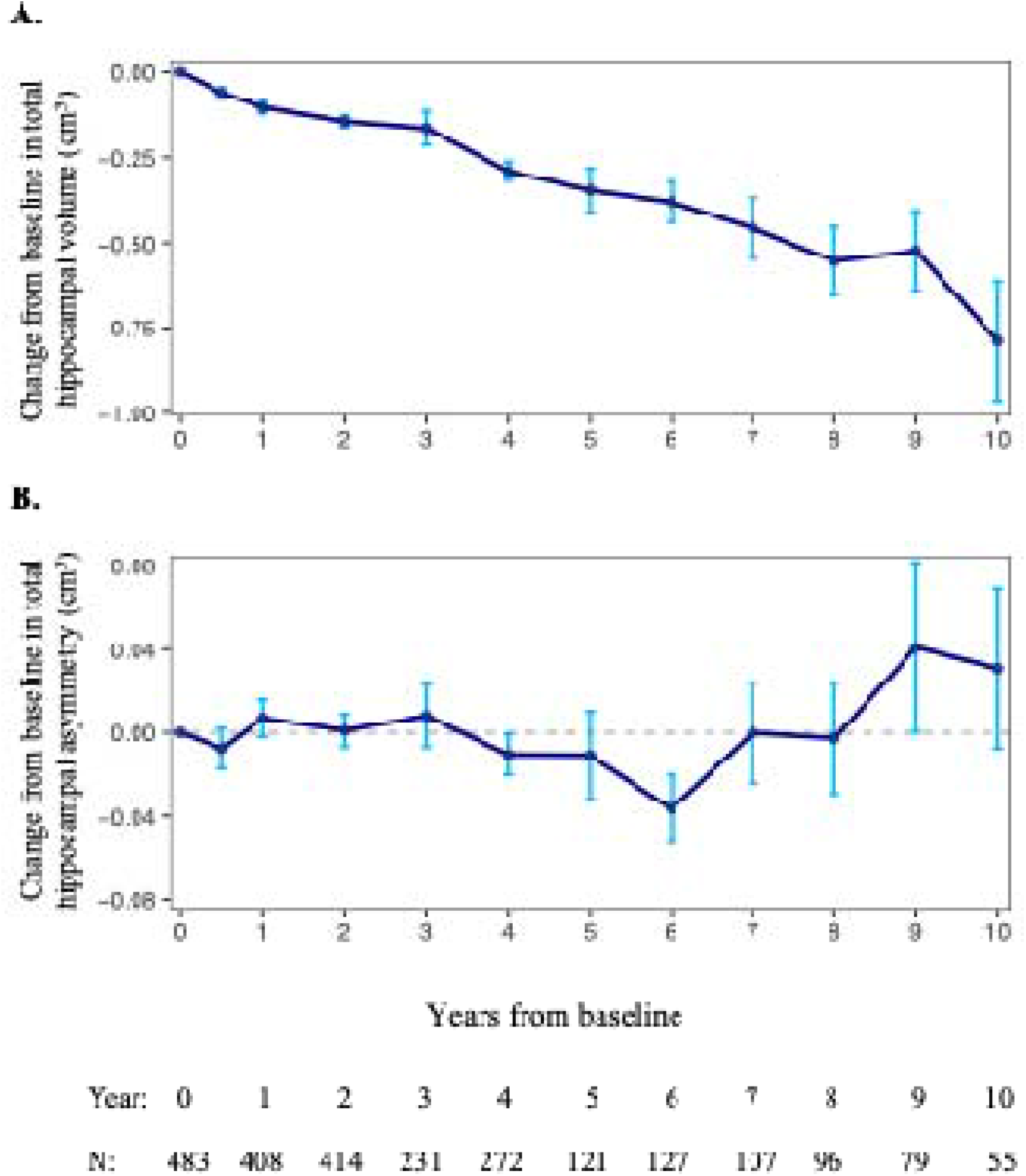
Mean change from baseline in total hippocampal volume (A) and hippocampal asymmetry (B) in the overall cohort across follow-up visits. For each participant, values at each visit were expressed as the difference from their baseline measurement. Points represent the mean across participants at each time point, and error bars indicate the standard error of the mean. Positive values indicate an increase relative to baseline, whereas negative values indicate a decrease.

### 3.2. Association of baseline p-tau181 with hippocampal asymmetry over time

In the base model, higher p-tau181 levels were significantly associated with faster increase in hippocampal asymmetry (β = 1.20 ± 0.43, *p* = 0.006, Table 2). These associations remained significant after adjustment for baseline total hippocampal volume, and further for baseline hippocampal volume and Aβ42 (both *p* = 0.006). Higher p-tau181 levels were significantly associated with a faster decline in hippocampal total volume (β = −1.87 ± 0.60, *p* = 0.002, Table 3). These associations remained significant after adjustment for Aβ42. No significant associations were found between p-tau181 and hippocampal asymmetry and total volume at baseline.

**Table 2.** Associations of CSF biomarkers with baseline hippocampal asymmetry and longitudinal change.

|  | p-tau181 |  |  |  | Aβ42 |  |  |  |
| --- | --- | --- | --- | --- | --- | --- | --- | --- |
|  | Baseline β (SE) | p | Longitudinal β (SE) | p | Baseline β (SE) | p | Longitudinal β (SE) | p |
| Model 1 | 0.08 (1.96) | 0.967 | 1.20 (0.43) | 0.006 | 0.01 (0.03) | 0.709 | 0.00 (0.01) | 0.752 |
| Model 2 | 0.04 (1.96) | 0.982 | 1.20 (0.43) | 0.006 | 0.01 (0.03) | 0.715 | 0.00 (0.01) | 0.751 |
| Model 3 | 0.22 (1.99) | 0.913 | 1.23 (0.43) | 0.006 | 0.01 (0.03) | 0.674 | 0.00 (0.01) | 0.546 |

Model 1 adjusted for age, sex, education, APOE ε4 status, time, the baseline CSF biomarker (p-tau181 or Aβ42), and the corresponding biomarker × time interaction.
Model 2 additionally adjusted for baseline total hippocampal volume.
Model 3 additionally adjusted for the other CSF biomarker (Aβ42 in p-tau181 models and p-tau181 in Aβ42 models).
Abbreviations: Aβ, amyloid-beta 42; p-tau, phosphorylated tau at threonine 181.

**Table 3.** Associations of CSF biomarkers with baseline total hippocampal volume and longitudinal change.

|  | p-tau181 |  |  |  | Aβ42 |  |  |  |
| --- | --- | --- | --- | --- | --- | --- | --- | --- |
|  | Baseline β (SE) | p | Longitudinal β (SE) | p | Baseline β (SE) | p | Longitudinal β (SE) | p |
| Model 1 | 2.13 (4.16) | 0.609 | -1.87 (0.60) | 0.002 | 0.04 (0.06) | 0.485 | -0.03 (0.01) | 0.002 |
| Model 2 | 2.54 (4.23) | 0.547 | -1.96 (0.59) | 0.001 | 0.04 (0.06) | 0.448 | -0.03 (0.01) | < 0.001 |
| Model 3 | 2.50 (4.23) | 0.556 | -1.96 (0.59) | 0.001 | 0.04 (0.06) | 0.451 | -0.03 (0.01) | < 0.001 |

Model 1 adjusted for age, sex, education, APOE ε4 status, time, the baseline CSF biomarker (p-tau181 or Aβ42), and the corresponding biomarker × time interaction.
Model 2 additionally adjusted for the alternate CSF biomarker.
Model 3 additionally adjusted for baseline hippocampal asymmetry.
Abbreviations: Aβ, amyloid-beta 42; p-tau, phosphorylated tau at threonine 181.

In contrast, there were no significant associations between baseline Aβ42 levels and hippocampal asymmetry over time (Table 2). However, higher baseline amyloid burden was significantly associated with a faster rate of total hippocampal atrophy over time (β = −0.03± 0.01, *p* = 0.002, Table 3). These associations remained significant after controlling for baseline p-tau181.

### 3.3. Effect of time on hippocampal atrophy and asymmetry

Linear mixed-effects models demonstrated a significant longitudinal decline in total hippocampal volume over time (β_{time} = −0.07, *p* < 0.001; Fig. 2A). In contrast, hippocampal asymmetry did not show a significant overall change over time (β_{time} = −0.02, *p* = 0.106; Fig. 2B), suggesting that left–right hippocampal volumetric differences remained relatively stable after adjusting for baseline covariates.

**Figure 2.**
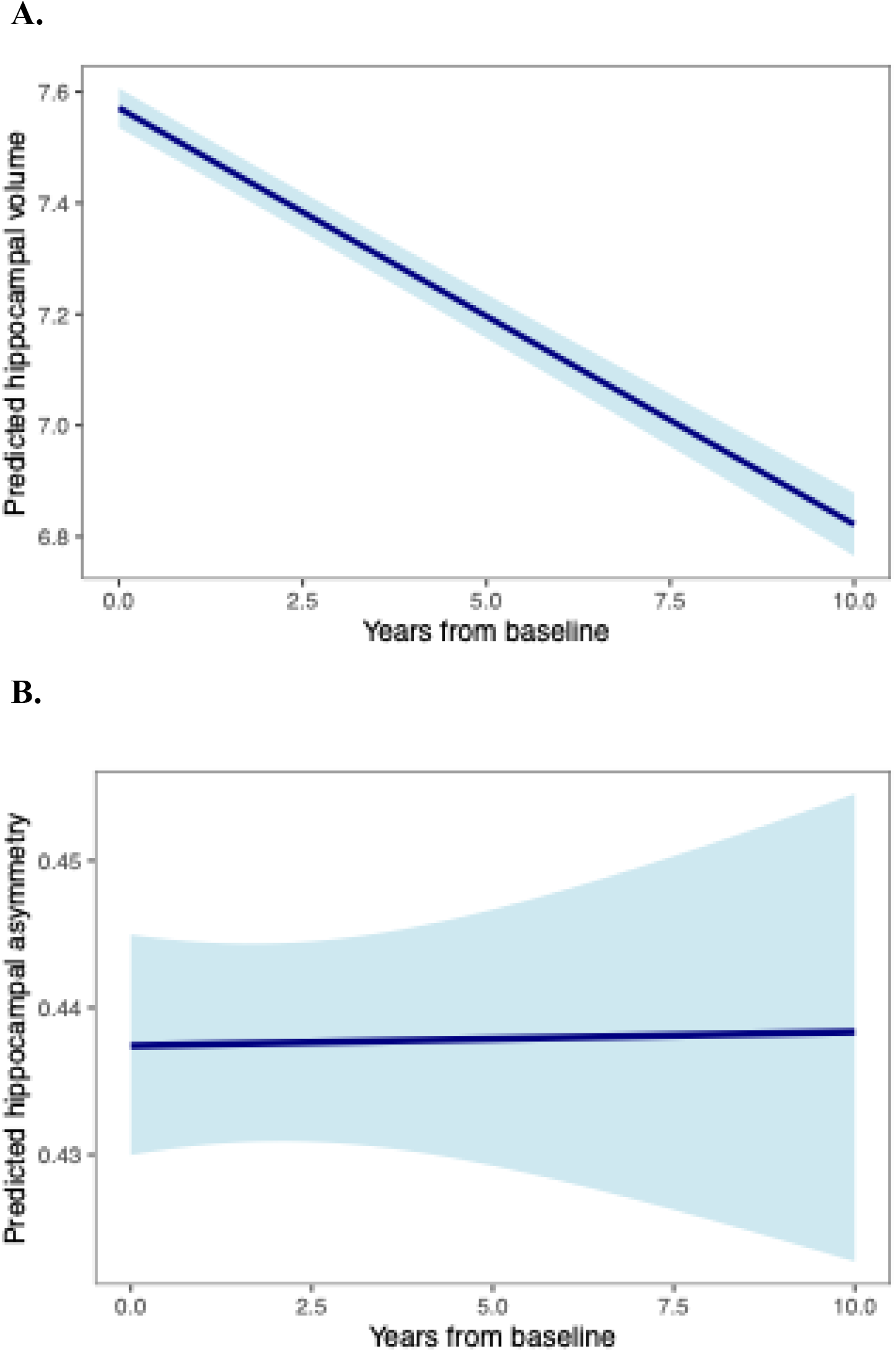
Model-predicted longitudinal trajectories of total hippocampal volume (A) and hippocampal asymmetry (B) over time. Lines represent estimated values from linear mixed-effects models. Shaded areas represent ±1 standard error of the mean. Models were adjusted for age, sex, education, APOE ε4 status, time, CSF biomarkers and their interactions with time; models of hippocampal asymmetry were additionally adjusted for baseline total hippocampal volume.

When stratified by baseline dichotomous p-tau181 status, model-predicted trajectories demonstrated divergent longitudinal patterns of hippocampal asymmetry (Fig. 3A). This divergence suggests that elevated baseline p-tau181 (> 27pg/mL) was associated with increasingly hippocampal asymmetry over time. However, when stratified by baseline amyloid status, model-predicted trajectories suggested modest differences in longitudinal patterns of hippocampal asymmetry (Fig. 3B), although the amyloid × time interaction was not significant.

**Figure 3.**
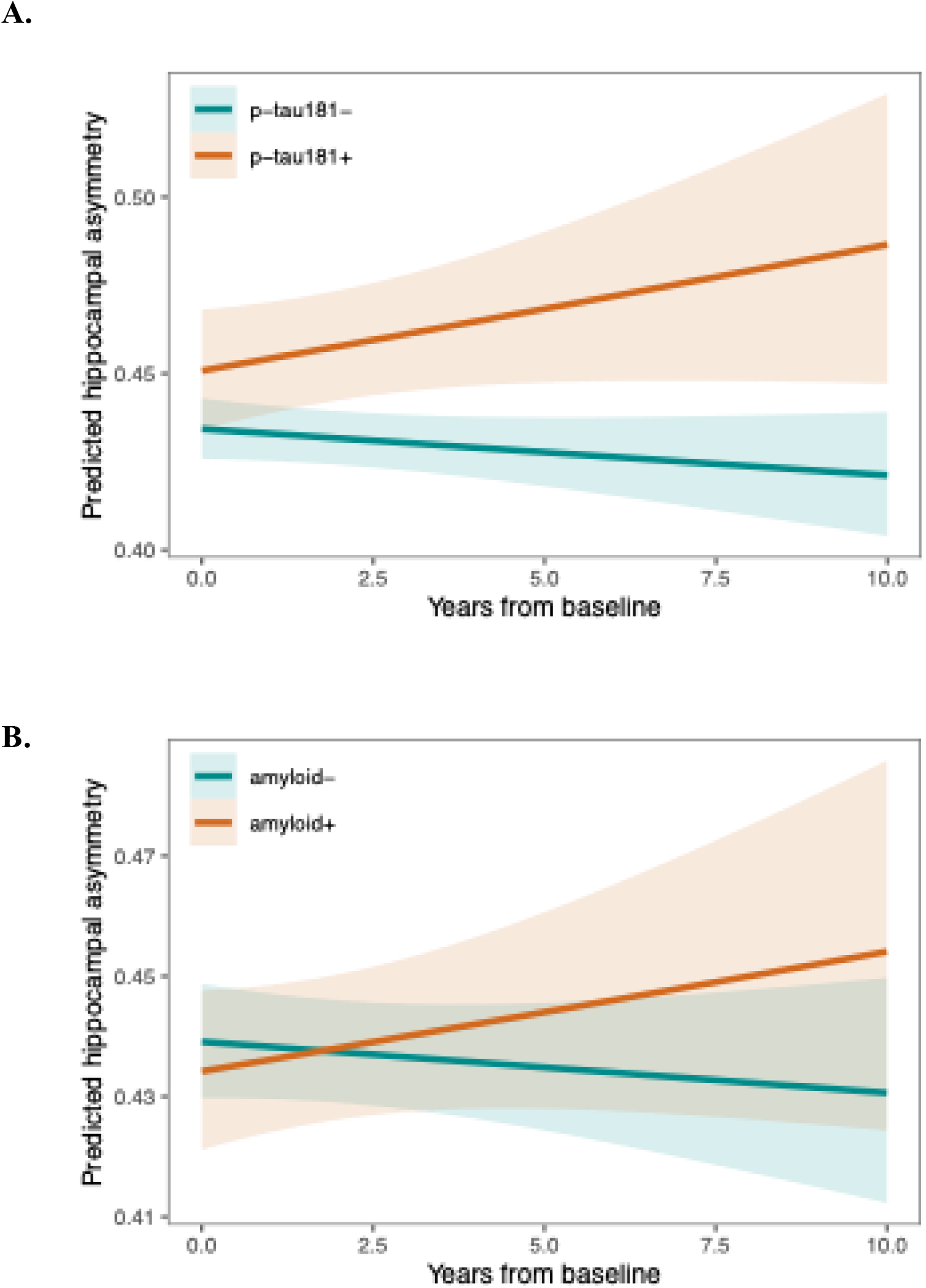
Model-predicted longitudinal trajectories of hippocampal asymmetry by binary baseline p-tau181 (**A**) and Aβ42 (**B**) status. Lines represent estimated values from linear mixed-effects models for participants with lower (0) and higher (1) levels of each biomarker at baseline. Shaded areas represent ±1 standard error of the mean. Model was adjusted for age, sex, education, APOE ε4 status, time, CSF biomarkers and their interactions with time, and baseline hippocampal volume.

In separate models fitted to evaluate the association between p-tau181 and longitudinal trajectories of left and right hippocampal volumes, p-tau181 was significantly associated with faster longitudinal decline in both hippocampi (β = −0.99, SE = 0.32, *p* = 0.002 for the left, and β = −0.80, SE = 0.33, *p* = 0.015 for the right).

### 3.4. Association of p-tau181 with hippocampal asymmetry and total volume stratified by amyloid status

Although the amyloid × time interaction was not statistically significant; to further investigate the effect of amyloid on the association of p-tau181 and hippocampal asymmetry, we conducted exploratory analyses by dichotomous Aβ status (Table 4). Higher p-tau181 levels were associated with faster increase in hippocampal asymmetry over time only among Aβ-individuals (β = 1.41, SE = 0.55, *p* = 0.014), while in the Aβ+ group no significant associations were observed. In contrast, higher p-tau181 levels were associated with faster total hippocampal volume atrophy only among Aβ+ individuals (β = −2.12, SE = 0.61, *p* < 0.001), and not in the Aβ-group.

**Table 4.**
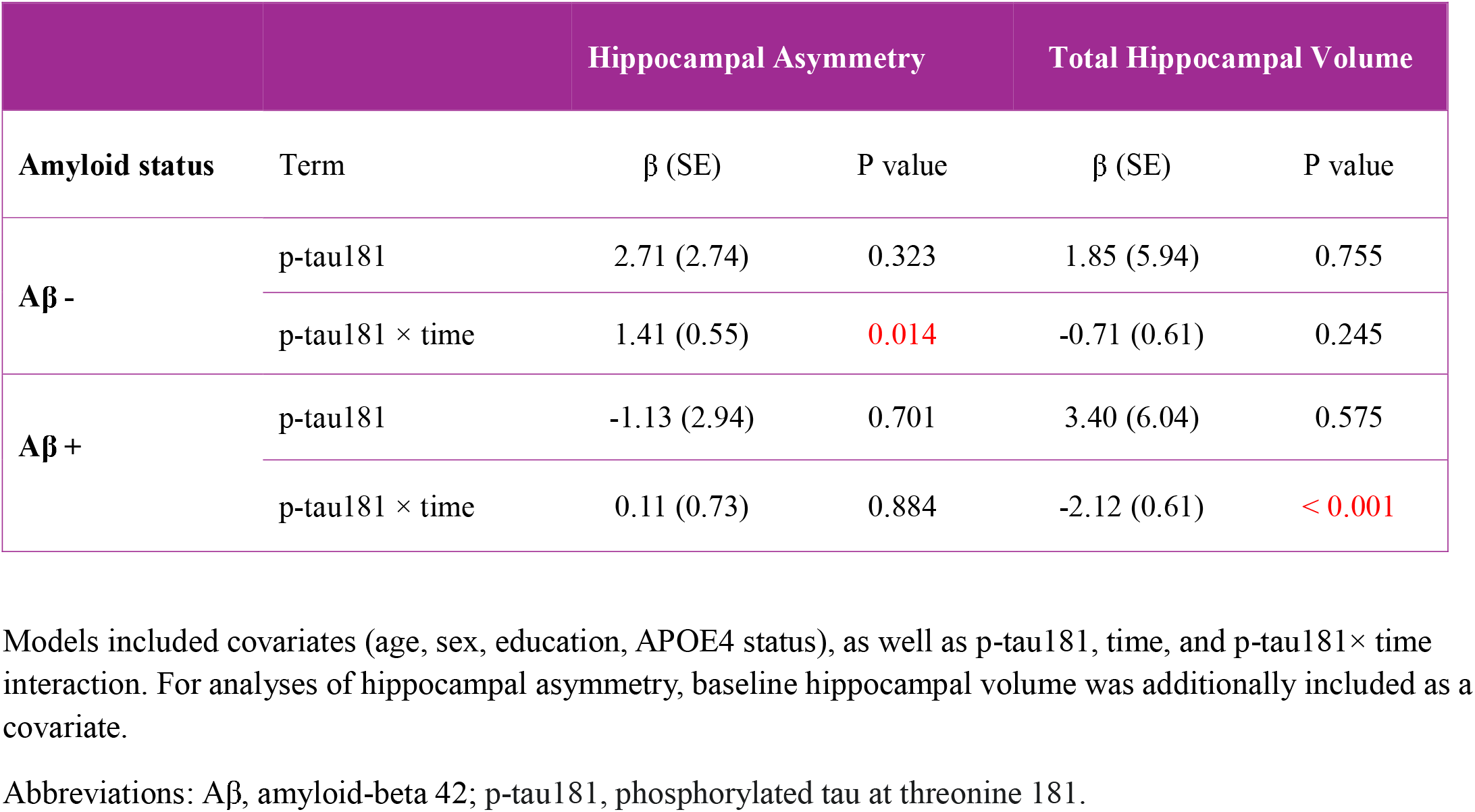
Association baseline CSF p-tau181 with hippocampal measures, stratified by baseline amyloid status.

### 3.5 Comparison of p-tau181, total tau, and tau PET

In this cohort, CSF p-tau181 and total tau (t-tau) were highly correlated (Spearman’s ρ = 0.98, *p* < 0.001). Consistent with this, the association of t-tau with longitudinal changes in hippocampal asymmetry (β = 0.11 ± 0.04, *p* = 0.015) was similar to that observed for p-tau181 (β = 1.23 ± 0.43, *p* = 0.006) in the full model including all covariates.

Among 130 of 483 participants with baseline tau PET, temporal meta-ROI SUVR was modestly and significantly correlated with CSF p-tau181 (Spearman’s ρ = 0.25, p = 0.004) and t-tau (Spearman’s ρ = 0.23, p = 0.009). After adjusting for age, sex, and CSF Aβ42, these associations were stronger, with partial correlation coefficient of r = 0.37 for p-tau181 and r = 0.33 for t-tau, both p < 0.001.

## 4. Discussion

In this longitudinal study of cognitively unimpaired individuals, we found that higher baseline CSF p-tau181 was associated with faster increase in hippocampal asymmetry over time, independent of baseline total hippocampal volume and amyloid burden. Higher baseline p-tau181 was also associated with increased total hippocampal atrophy, confirming its established relationship with neurodegeneration. In contrast, higher amyloid burden was associated with longitudinal decline in total hippocampal volume but not with change in hippocampal asymmetry. In stratified analyses, the association between p-tau181 and increasing asymmetry remained significant in Aβ− individuals, while the association between p-tau181 and faster total hippocampal atrophy was significant in Aβ+ individuals. Together, these findings suggest that CSF p-tau181 is associated not only with overall hippocampal tissue loss but also with the pattern by which that tissue loss unfolds across hemispheres, and that these two dimensions of neurodegeneration may relate differently to amyloid status.

These findings position hippocampal asymmetry as more than a secondary descriptor of hippocampal atrophy. Total hippocampal volume is a bilateral summary measure of neurodegenerative burden, whereas asymmetry captures left-right imbalance and therefore may index selective or lateralized vulnerability within the MTL. That distinction is important because two individuals may have similar total hippocampal volumes but differ substantially in how degeneration is distributed across hemispheres. Our data suggest that this lateralized component is not simply noise around the global atrophy signal but may represent a biologically meaningful dimension of neurodegeneration. Prior literature on hippocampal asymmetry in AD has been mixed, with some studies reporting limited evidence for disease-related asymmetry and more recent synthesis suggesting greater left-sided volume reduction across AD and MCI groups, underscoring that asymmetry is not a uniform feature of disease severity and may instead reflect subtype- or pathology-specific processes.^10^

In a recent work, we have shown that while total hippocampal volume was associated with cognitive performance in multiple domains, greater hippocampal asymmetry was mainly associated with accelerated memory decline, especially among Aβ-individuals.^15^ That prior observation is important here because it suggests that asymmetry is not merely an alternative structural summary, but a phenotype with distinct cognitive correlates. The current findings extend that work by linking asymmetric degeneration specifically to p-tau181.

Previous studies have shown that spatially and temporally, tau pathology correlates much more strongly than amyloid with neurodegeneration and cognitive impairment.^23^ It has been shown that tau burden had both direct and atrophy-mediated effects on cognition, and these associations were weakly related to amyloid burden.^24^ Although tau pathology is best assessed using imaging modalities, several studies have demonstrated that CSF p-tau181 levels closely reflect cerebral tau burden.^25–27^

The link between CSF p-tau181 and global hippocampal atrophy has been previously reported.^28^ In a well-defined AD cohort, CSF total tau and p-tau181 were inversely associated with hippocampal volume, whereas CSF Aβ42 was not associated with any brain measure, consistent with neurodegeneration driven by tau pathology in AD.^29^ In ADNI, CSF p-tau181 predicts faster hippocampal atrophy particularly among individuals with mild cognitive impairment and AD,^30^ and p-tau181 demonstrated the strongest association with hippocampal atrophy, whereas Aβ42 showed the weakest.^31^ Similarly, an association between CSF p-tau181and accelerated hippocampal atrophy has been demonstrated in cognitively normal older adults in ADNI.^32^ Our findings extend this literature in two ways. First, they confirm in cognitively unimpaired individuals that p-tau181 is associated with faster decline in total hippocampal volume even after accounting for amyloid. Second, they suggest that tau-related neurodegeneration may not be fully captured by total hippocampal volume alone, because p-tau181 was also associated with increasing hippocampal asymmetry over time.

Importantly, our findings position hippocampal asymmetry as a distinct and potentially informative neuroimaging phenotype. The observation that asymmetry remained relatively stable at the group level but diverged according to baseline p-tau181 levels suggests that asymmetric atrophy is not simply a byproduct of global atrophy, but rather reflects an individualized trajectory influenced by underlying pathology.

The differential patterns observed in amyloid-stratified analyses provide further information on the underlying biological mechanisms of p-tau-asymmetry associations. However, these subgroup findings should be interpreted cautiously. Because the statistical significance of an association in one subgroup and not in another does not by itself establish a significant between-group difference, the present results should be viewed as suggestive rather than definitive evidence of effect modification by amyloid status. Even so, the pattern is biologically interesting as p-tau181 was associated with increasing hippocampal asymmetry in Aβ-individuals, and with total hippocampal atrophy in Aβ+ individuals. One possible interpretation is that in Aβ+ individuals, p-tau181 reflects a more canonical AD-related neurodegenerative process expressed primarily as overall hippocampal tissue loss, whereas in Aβ− individuals p-tau181 may be indexing a more regionally selective medial temporal process that manifests as increasing asymmetry. This interpretation is consistent with recent longitudinal works showing the association of tau with hippocampal atrophy, independent of amyloid, in cognitively normal older adults and different downstream structural pathways for tau in Aβ− versus Aβ+ individuals.^33, 34^

One potential explanation for our findings is the contribution of other age-related co-pathologies that preferentially affect medial temporal structures. Conditions such as primary age-related tauopathy (PART) are common in cognitively unimpaired older adults and is characterized by tau accumulation in the MTL in the absence of significant amyloid pathology.^35–37^ Similarly, LATE and hippocampal sclerosis of aging often co-occur with tau pathology in older adults and are characterized by MTL-predominant neurodegeneration that may be disproportionate or asymmetric.^38, 39^ Late-life hippocampal sclerosis, in particular, is often asymmetric or unilateral at autopsy, and has been associated with disproportionately severe hippocampal atrophy.^40^ Although we do not have in-vivo measures to quantity these pathologies, the observed association between p-tau181 and asymmetric atrophy in Aβ-individuals is compatible with the possibility that hippocampal asymmetry reflects non-amyloid predominant MTL neurodegeneration, potentially arising from mixed or overlapping age-related pathologies rather than AD neuropathologic change alone.

This study has several strengths. Simultaneous modeling of total hippocampal volume and asymmetry allows direct comparison of two related but potentially distinct structural expressions of neurodegeneration. In addition, the focus on cognitively unimpaired individuals enhances the relevance of these findings to the preclinical stages of AD. Identifying imaging markers that capture early and heterogeneous neurodegenerative processes is critical for improving risk stratification and refining participant selection in clinical trials. Furthermore, the integration of CSF biomarkers with repeated MRI measures over extended follow-up enables a more comprehensive characterization of longitudinal neurodegenerative trajectories. A few limitations should be considered. First, CSF biomarkers were measured only at baseline, limiting our ability to assess longitudinal changes in tau and amyloid. Second, the absence of in vivo markers of non-AD pathologies, such as TDP-43, precludes direct attribution of asymmetric atrophy to specific underlying mechanisms. Finally, the ADNI cohort is a highly selected predominantly White, highly educated, and with relatively few comorbidities, thus it is not fully representative of the general population, potentially limiting the generalizability of results.

In conclusion, baseline CSF p-tau181 was associated with both faster total hippocampal atrophy and increasing hippocampal asymmetry over time, whereas amyloid burden was associated with overall hippocampal atrophy but not with asymmetry. These results support the idea that the magnitude and the lateralization of hippocampal neurodegeneration may reflect partially distinct biological processes. Measures of hippocampal asymmetry may therefore add information beyond total hippocampal volume, particularly in cognitively unimpaired older adults in whom non-amyloid-predominant medial temporal degeneration may already be emerging. Incorporating measures of hemispheric asymmetry into neuroimaging frameworks could improve characterization of early and heterogeneous pathways of neurodegeneration in aging and AD.

## Data Availability

All data produced in the present study are available upon reasonable request to the authors

https://adni.loni.usc.edu/

## Acknowledgment

Data collection and sharing for this project was funded by the Alzheimer’s Disease Neuroimaging Initiative (ADNI) (National Institutes of Health Grant U01 AG024904) and DOD ADNI (Department of Defense award number W81XWH-12-2-0012). ADNI is funded by the National Institute on Aging, the National Institute of Biomedical Imaging and Bioengineering, and through generous contributions from the following: AbbVie, Alzheimer’s Association; Alzheimer’s Drug Discovery Foundation; Araclon Biotech; BioClinica, Inc.; Biogen; Bristol-Myers Squibb Company; CereSpir, Inc.; Cogstate; Eisai Inc.; Elan Pharmaceuticals, Inc.; Eli Lilly and Company; EuroImmun; F. Hoffmann-La Roche Ltd and its affiliated company Genentech, Inc.; Fujirebio; GE Healthcare; IXICO Ltd.; Janssen Alzheimer Immunotherapy Research & Development, LLC.; Johnson & Johnson Pharmaceutical Research & Development LLC.; Lumosity; Lundbeck; Merck & Co., Inc.; Meso Scale Diagnostics, LLC.; NeuroRx Research; Neurotrack Technologies; Novartis Pharmaceuticals Corporation; Pfizer Inc.; Piramal Imaging; Servier; Takeda Pharmaceutical Company; and Transition Therapeutics. The Canadian Institutes of Health Research is providing funds to support ADNI clinical sites in Canada. Private sector contributions are facilitated by the Foundation for the National Institutes of Health (www.fnih.org). The grantee organization is the Northern California Institute for Research and Education, and the study is coordinated by the Alzheimer’s Therapeutic Research Institute at the University of Southern California. ADNI data are disseminated by the Laboratory for Neuro Imaging at the University of Southern California.

